# How Efficacious Must a COVID-19 Coronavirus Vaccine be to Prevent or Stop an Epidemic by Itself

**DOI:** 10.1101/2020.05.29.20117184

**Authors:** Sarah M. Bartsch, Kelly J. O’Shea, Marie C. Ferguson, Maria Elena Bottazzi, Sarah N. Cox, Ulrich Strych, James A. McKinnell, Patrick T. Wedlock, Sheryl S. Siegmund, Peter J. Hotez, Bruce Y. Lee

## Abstract

**Background:** Given the continuing coronavirus disease 2019 (COVID-19) pandemic and much of the U.S. implementing social distancing due to the lack of alternatives, there has been a push to develop a vaccine to eliminate the need for social distancing.

**Methods:** In 2020, we developed a computational model of the U.S. simulating the spread of COVID-19 coronavirus and vaccination.

**Results:** Simulation experiments revealed that when vaccine efficacy exceeded 70%, coverage exceeded 60%, and vaccination occurred on day 1, the attack rate dropped to 22% with daily cases not exceeding 3.2 million (reproductive rate, R_0_, 2.5). When R_0_ was 3.5, the attack rate dropped to 41% with daily cases not exceeding 14.4 million. Increasing coverage to 75% when vaccination occurred by day 90 resulted in 5% attack rate and daily cases not exceeding 258,029when R_0_ was 2.5 and a 26% attack rate and maximum daily cases of 22.6 million when R_0_ was 3.5. When vaccination did not occur until day 180, coverage (i.e., those vaccinated plus those otherwise immune) had to reach 100%. A vaccine with an efficacy between 40% and 70% could still obviate the need for other measures under certain circumstances such as much higher, and in some cases, potentially unachievable, vaccination coverages.

**Conclusion:** Our study found that to either prevent or largely extinguish an epidemic without any other measures (e.g., social distancing), the vaccine has to have an efficacy of at least 70%.

## Introduction

With the continuing coronavirus disease 2019 (COVID-19) pandemic and much of the U.S. having implemented social distancing measures due to the lack of alternatives, there has been a push for efforts to develop a vaccine to eliminate the need for social distancing measures. However, as these vaccine development efforts progress, it is not yet clear what vaccine efficacies are necessary to achieve this goal. As we have described previously,^1, 2^ it is important to determine efficacy thresholds to aim for early on and during a vaccine’s development. This is especially true in early-stage development when changes can still be implemented more easily. Currently, there are sixteen vaccine candidates are under clinical phase I-II evaluation and over 100 preclinical candidate vaccines are in the immunization pipeline.^3–6^ In order to help establish the ideal goals for the vaccine efficacies needed to prevent and extinguish a COVID-19 coronavirus epidemic, we developed a computational simulation model representing the United States population, the spread of SARS-CoV-2, and the impact of a vaccine under various conditions.

## Methods

### Model Structure

Using Microsoft Excel (Microsoft Corporation, Redmond, WA) with the Crystal Ball add-in (Oracle Corporation, Redwood Shore, CA), we developed a computational model (in 2020) representing the U.S. population (357,157,434 persons) and their different interactions with each other as well as the spread of SARS-CoV-2 and the potential health and economic outcomes^7^. The model advances in discrete, one-day time steps for 2.5 years (which is the longest duration of the epidemic in the evaluated scenarios included in our study). On any given day, each individual in the model is in one of five mutually exclusive SARS-CoV-2 states: 1) susceptible (S, not infected and able to become infected), 2) exposed (E, infected, but not able to transmit to others), 3) infectious and asymptomatic (I_a_, infected, but without symptoms, and able to transmit to others), 4) infectious and symptomatic (I_s_, infected, showing symptoms, and able to transmit to others), or 5) recovered/immune (R, not infected and unable to become infected). On day one, a set number of individuals start in the ‘Ia state’ and ‘Is state’ (i.e., seed coronavirus), with the remainder starting in the ‘S state’. Each day, individuals interact with each other, and a person that is infectious can potentially transmit the virus to a person who is susceptible. If a susceptible person comes in contact with an infectious person, he/she moves from the ‘S state’ to the ‘E state.’ The following equation determines the number of susceptible individuals who became exposed each day: β^*^S^*^Is + (β^*^0.5)^*^S^*^Ia. Beta (β) equals the basic reproduction number (R_0_; the average number of secondary cases generated by one infectious case) divided by the infectious period duration and the number of individuals in the population, ‘S’ and ‘I’ represent the number of susceptible and infectious persons, respectively, on any given day. Exposed individuals remain in the ‘E state’ for the latent period duration (i.e., time between exposure and ability to transmit) before becoming infectious and moving to the ‘I state’ (at a rate of 1/latent period duration). As individuals can transmit the virus prior to disease onset^8^, we assume they could transmit one day prior to the start of symptoms. Each individual moving to the ‘I state’ has a probability of being symptomatic, which governs if they are in the ‘I_s_’ or ‘I_a_ state.’ Each person in the model draws an infectious period duration from a distribution (range: 4–15 days, including the day prior to symptom onset). Infectious individuals remain in the ‘I state’ until they recover and are no longer infectious, moving from the ‘I state’ to the ‘R state’ (at the rate of 1/infectious period duration).

Vaccination occurs on different days during the epidemic (varying with scenario) and protects in two different ways: by preventing infection and by preventing symptomatic disease (i.e., reducing viral shedding). Individuals who are vaccinated move into the ‘V state’. Each day, vaccinated individuals mix with others and if they come in contact with an infectious person, they move to the ‘E state’. When the vaccine prevents infection, individuals in the ‘E state’ will move to the ‘R state’ based on the vaccine efficacy. (Individuals for whom the vaccine is not effective move to either the ‘Ia state’ or ‘Is state’.) When the vaccine reduces viral shedding, it reduces their probability of moving to the ‘Is state’ based on the vaccine efficacy. Effectively vaccinated individuals in the ‘E state’ move to the ‘Ia state’, where they transmit at a lower rate. We assume vaccination protection to be immediate (i.e., on day of vaccination) and that the vaccine had no impact on individuals who had already been infected and/or exposed to SARSCoV-2.

Each symptomatically infected person (i.e., COVID-19 case) travels through a probability tree of different sequential outcomes.^7^ An infected person showing symptoms starts with a mild infection and has a probability of seeking ambulatory care or calling his/her physician (i.e., telephone consult). This person then has a probability of progressing to severe disease requiring hospitalization. If this person has only mild illness and is not hospitalized, he/she self-treats with over-the-counter medications and misses school or work for the duration of symptoms. If this person is hospitalized, he/she has a probability of developing severe pneumonia or severe non-pneumonia symptoms and has a probability of intensive care unit (ICU) admission. This patient then has a probability of having either sepsis or acute respiratory distress syndrome (ARDS), with or without sepsis. If this patient has ARDS, he/she requires the use of a ventilator. If the person is hospitalized, he/she has a probability of dying from COVID-19 complications. The person accrues relevant costs and health effects as he/she travels through the model.

The third-party payer perspective includes direct costs (e.g., ambulatory care, hospitalization), while the societal perspective includes direct and indirect (i.e., productivity losses due to absenteeism and mortality) costs. Hourly wage across all occupations^9^ serves as a proxy for productivity losses. Absenteeism results in productivity losses for the duration of symptoms. Death results in the net present value of productivity losses for missed lifetime earnings based on annual wage^9^ for the years of life lost based on an individual’s life expectancy.^10^

### Data Sources

Appendix Table 1 shows key model input parameters, values, and sources. All costs, clinical probabilities, and durations were age-specific when available and come from scientific literature or nationally representative data sources. Age-specific COVID-19 data are specific to the US context as of March 16, 2020.^11^ We report all costs in 2020 values, discounting all past and future values using a 3% rate. We parameterized seeding SARS-CoV2-infected persons into the population for a given R_0_ such that simulated cases reflected case data reported as of March 24, 2020.^11^

### Modeled Scenarios and Sensitivity Analyses

We evaluated the impact of introducing a vaccine with varying efficacies in the absence of other measures from the third-party payer and societal perspectives. Experiments consisted of 1,000 trial Monte Carlo simulations, varying parameters throughout their range (Appendix Table 1). Our initial scenario assumes an unmitigated epidemic with no vaccination. Experimental scenarios consisted of vaccinating individuals to either prevent infection or prevent symptomatic disease. Sensitivity analyses varied the vaccine efficacy (20%-100%), population coverage (those vaccinated and those otherwise immune, 50%-100%), timing of vaccination (0–180 days from the epidemic start), and R_0_ (2.5–3.5).^12–14^ We report results as the median and 95% uncertainty interval (UI).

## Results

### No Vaccine

Figures 1–2 (and Appendix Figure 1) show the resulting number of persons infected with SARSCoV-2 over time (i.e., epidemic curves) when the R_0_ was 2.5 and 3.5 in the absence of vaccination, while Table 1 (and Appendix Table 2) show the number of cases, clinical outcomes, resources used, and costs over the duration of the epidemic. When the R_0_ of the virus was 2.5, the total attack rate was 86% with a maximum number of daily SARS-CoV-2 infections (i.e., epidemic peak) of 45.0 million. A higher R_0_ of 3.5, resulted in a total attack rate of 96% with 72.7 million cases at the epidemic peak.

**Figure 1.**
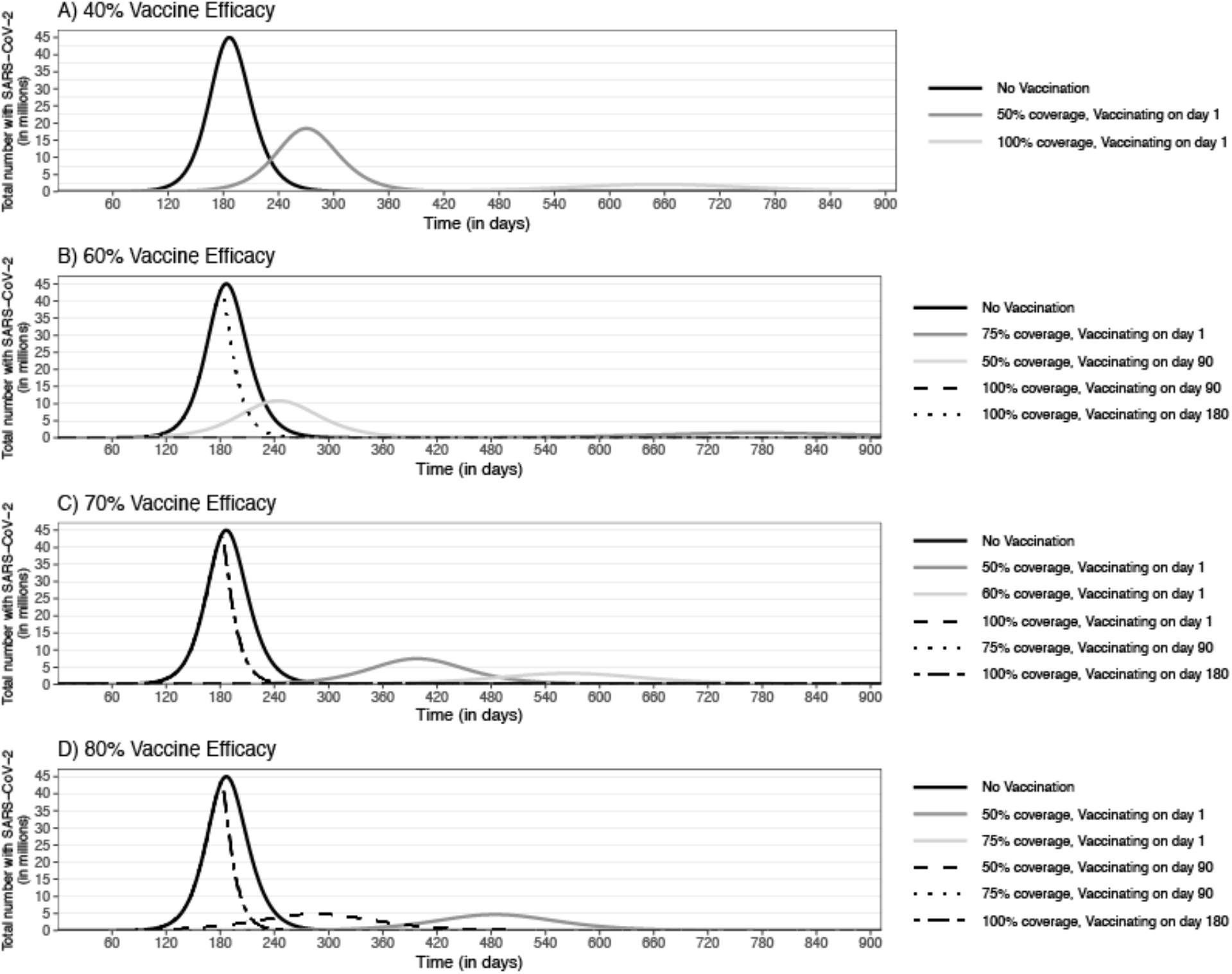
Epidemic curves for an epidemic with an R0 of 2.5 and the impact of a vaccine that prevents infection implemented at different points in the epidemic with a vaccine efficacy of A) 40%, B) 60%, C) 70%, and D) 80%

**Figure 2.**
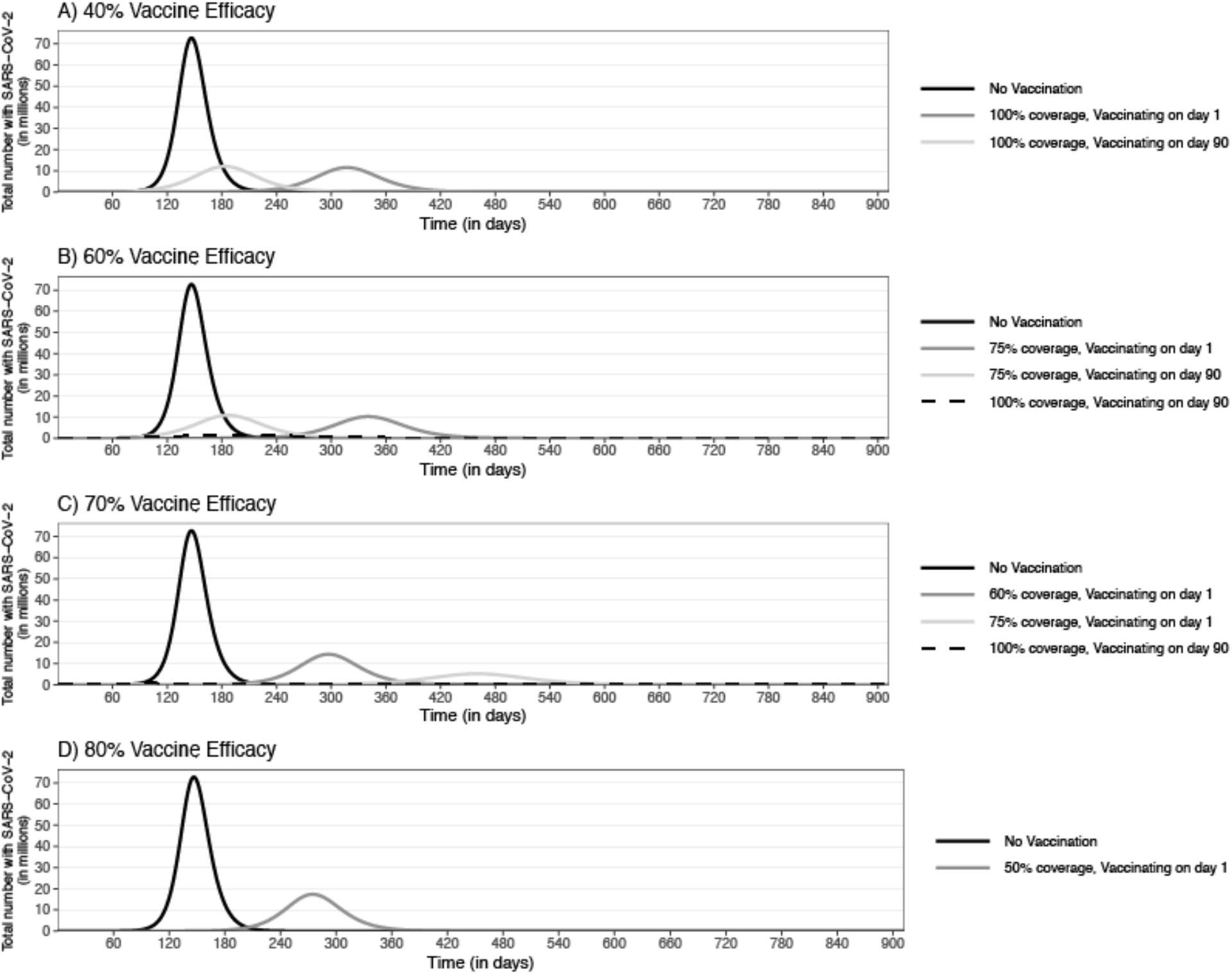
Epidemic curves for an epidemic with an R0 of 3.5 and the impact of a vaccine that prevents infection implemented at different points in the epidemic with a vaccine efficacy of A) 40%, B) 60%, C) 70%, and D) 80%

**Table 1.** Number of clinical outcomes, resource use, and costs [median (95% uncertainty interval)] due to COVID-19 during the course of an epidemic and the impact of vaccination with a vaccine that prevents infection, varying with vaccine efficacy

|  | Total Infected<br>with SARS-CoV-2 (in<br>millions) | Symptomatic<br>Cases<br>(in millions) | Hospitalized<br>(in millions) | Number of<br>Patients<br>Ventilated<br>(in millions) | Deaths<br>(in thousands) | Total Beds<br>Days<br>(in millions) | Ventilated<br>Days<br>(in millions) | Direct Medical<br>Costs<br>(in billions) | Productivity Losses<br>(in billions) |
| --- | --- | --- | --- | --- | --- | --- | --- | --- | --- |
| **R0 of 2.5** |  |  |  |  |  |  |  |  |  |
| No Vaccination | 282.5<br>(280.7-284.3) | 232.1<br>(223.9-240.0) | 48.1<br>(46.4-49.8) | 8.5<br>(6.5-9.7) | 4,160.9<br>(4,014.2-4,302.0) | 267.2<br>(251.7-282.6) | 68.0<br>(51.8-77.9) | 883.5<br>(808.5-980.8) | 2,796.4<br>(1,661.8-4,515.5) |
| **Vaccine Efficacy 40%** |  |  |  |  |  |  |  |  |  |
| 75% coverage,<br>vaccinating on day 1 | 121.7<br>(113.1-131.8) | 109.1<br>(101.9-116.9) | 22.6<br>(21.1-24.2) | 4.0<br>(3.0-4.6) | 1,955.5<br>(1,826.8-2,095.2) | 125.5<br>(115.8-136.5) | 32.0<br>(24.0-37.2) | 411.1<br>(367.0-459.8) | 1,293.8<br>(763.9-2,098.5) |
| 100% coverage,<br>vaccinating on day 1 | 55.6<br>(26.1-66.2) | 54.3<br>(25.3-64.5) | 11.3<br>(5.2-13.4) | 1.9<br>(0.9-2.5) | 974.2<br>(453.6-1,156.6) | 62.5<br>(29.4-74.6) | 15.5<br>(7.2-19.9) | 198.4<br>(89.4-244.6) | 594.7<br>(247.4-1,060.4) |
| **Vaccine Efficacy 60%** |  |  |  |  |  |  |  |  |  |
| 50% coverage,<br>vaccinating on day 1 | 138.8<br>(131.3-148.3) | 115.5<br>(109.8-122.3) | 24.0<br>(22.8-25.4) | 4.2<br>(3.3-4.9) | 2,071.1<br>(1,969.2-2,192.7) | 132.9<br>(123.6-142.7) | 33.8<br>(26.1-39.2) | 437.2<br>(389.2-488.9) | 1,408.2<br>(796.9-2,275.6) |
| 75% coverage,<br>vaccinating on day 1 | 43.0<br>(7.1-53.8) | 37.7<br>(6.2-48.5) | 7.8<br>(1.3-10.0) | 1.3<br>(0.2-1.9) | 675.9<br>(111.3-869.0) | 43.3<br>(7.2-56.1) | 10.5<br>(1.7-15.1) | 137.7<br>(22.5-183.9) | 386.6<br>(62.5-771.4) |
| 100% coverage,<br>vaccinating on day 90 | 1.7<br>(0.3-52.0) | 1.6<br>(0.3-47.5) | 0.3<br>(0.1-9.8) | 0.059<br>(0.011-1.7) | 29.1<br>(5.8-850.8) | 1.8<br>(0.4-54.2) | 0.5<br>(0.1-13.7) | 6.1<br>(1.2-176.9) | 20.1<br>(3.5-584.4) |
| **Vaccine Efficacy 70%** |  |  |  |  |  |  |  |  |  |
| 75% coverage,<br>vaccinating on day 90 | 14.9<br>(3.8-71.6) | 13.0<br>(3.2-61.5) | 2.7<br>(0.7-12.8) | 0.463<br>(0.110-2.318) | 232.6<br>(57.8-1,102.9) | 15.0<br>(3.7-71.1) | 3.7<br>(.9-18.2) | 48.6<br>(12.0-243.5) | 153.1<br>(34.3-805.0) |
| 100% coverage,<br>vaccinating on day 90 | 1.1<br>(0.2-34.5) | 1.0<br>(0.2-31.0) | 0.20<br>(0.04-6.43) | 0.035<br>(0.006-1.088) | 17.2<br>(3.3-555.6) | 1.1<br>(0.2-35.7) | 0.3<br>(0.051-8.8) | 3.7<br>(0.7-119.4) | 11.6<br>(1.9-359.2) |
| 100% coverage,<br>vaccinating on day 180 | 188.9<br>(22.1-283.8) | 158.7<br>(19.4-237.3) | 32.9<br>(4.0-49.2) | 5.7<br>(0.7-9.4) | 2,844.6<br>(348.7-4,254.7) | 181.7<br>(22.7-275.2) | 45.2<br>(5.5-75.2) | 605.9<br>(76.8-949.8) | 1,758.9<br>(221.5-4,103.8) |
| **Vaccine Efficacy 80%** |  |  |  |  |  |  |  |  |  |
| 75% coverage,<br>vaccinating on day 90 | 2.7<br>(0.5-59.5) | 2.3<br>(0.4-51.0) | 0.5<br>(0.1-10.6) | 0.084<br>(0.016-1.80) | 41.5<br>(7.9-914.2) | 2.7<br>(0.5-58.0) | 0.7<br>(0.1-14.3) | 8.8<br>(1.7-188.2) | 27.9<br>(4.9-660.1) |
| 100% coverage,<br>vaccinating on day 90 | 1.0<br>(0.2-35.2) | 0.8<br>(0.1-30.2) | 0.2<br>(0.027-6.3) | 0.030<br>(0.005-1.1) | 15.0<br>(2.4-541.3) | 1.0<br>(0.2-34.8) | 0.2<br>(0.04-8.7) | 3.2<br>(0.5-117.7) | 10.3<br>(1.5-331.9) |
| 100% coverage,<br>vaccinating on day 180 | 173.0<br>(15.6-283.6) | 142.7<br>(13.5-236.5) | 29.6<br>(2.8-49.0) | 5.0<br>(0.5-9.3) | 2,558.0<br>(241.3-4,240.2) | 164.3<br>(15.5-274.6) | 40.9<br>(4.0-74.2) | 556.4<br>(51.7-936.9) | 1,617.6<br>(144.4-3,944.4) |
| **Vaccine Efficacy 100%** |  |  |  |  |  |  |  |  |  |
| 75% coverage,<br>vaccinating on day 90 | 1.2<br>(0.2-36.8) | 0.9<br>(0.2-30.6) | 0.2<br>(0.03-6.3) | 0.034<br>(0.005-1.07) | 17.0<br>(2.7-548.6) | 1.1<br>(0.2-35.1) | 0.3<br>(0.043-8.5) | 3.6<br>(0.6-112.8) | 11.3<br>(1.6-354.3) |
| 75% coverage,<br>vaccinating on day 180 | 194.7<br>(20.7-283.6) | 160.3<br>(17.0-236.8) | 33.2<br>(3.5-49.1) | 5.6<br>(0.6-9.4) | 2,873.6<br>(304.8-4,244.7) | 183.8<br>(19.6-275.3) | 45.0<br>(4.9-75.0) | 600.6<br>(65.5-937.4) | 1,755.2<br>(180.8-4,028.3) |
| **R0 of 3.5** |  |  |  |  |  |  |  |  |  |
| No Vaccination | 312.9<br>(311.9-314.0) | 257.1<br>(248.7-264.8) | 53.3<br>(51.6-54.9) | 9.5<br>(7.2-10.7) | 4,608.3<br>(4,458.0-4,747.6) | 296.0<br>(281.4-312.1) | 76.2<br>(57.6-85.7) | 978.9<br>(896.4-1,082.3) | 3,141.3<br>(1,871.3-5,059.2) |
| **Vaccine Efficacy 40%** |  |  |  |  |  |  |  |  |  |
| 100% coverage,<br>vaccinating on day 1 | 118.5<br>(109.0-131.7) | 113.8<br>(106.9-120.3) | 23.6<br>(22.2-24.9) | 4.2<br>(3.1-4.8) | 2,041.0<br>(1,916.7-2,155.9) | 130.8<br>(121.5-140.2) | 33.5<br>(25.1-38.4) | 430.6<br>(384.3-482.6) | 1,348.2<br>(807.1-2,192.4) |
| **Vaccine Efficacy 60%** |  |  |  |  |  |  |  |  |  |
| 75% coverage,<br>vaccinating on day 1 | 113.5<br>(106.7-123.9) | 99.7<br>(94.5-105.5) | 20.7<br>(19.6-21.9) | 3.6<br>(2.8-4.2) | 1,787.1<br>(1,694.1-1,890.5) | 114.7<br>(107.0-123.0) | 29.2<br>(22.3-33.7) | 376.0<br>(337.3-420.8) | 1,201.7<br>(701.5-1,932.6) |
| 100% coverage,<br>vaccinating on day 1 | 4.7<br>(0.2-31.1) | 4.7<br>(0.2-30.7) | 0.965<br>(0.047-6.4) | 0.2<br>(0.008-1.13) | 83.5<br>(4.0-551.0) | 5.3<br>(0.3-35.3) | 1.4<br>(0.1-9.1) | 16.8<br>(0.8-114.8) | 51.6<br>(2.6-403.1) |
| 100% coverage,<br>vaccinating on day 90 | 35.5<br>(22.8-247.4) | 34.4<br>(21.8-208.0) | 7.1<br>(4.5-43.1) | 1.3<br>(0.8-7.5) | 616.3<br>(390.9-3,728.6) | 39.5<br>(25.1-240.0) | 10.1<br>(6.0-60.2) | 132.3<br>(81.6-788.0) | 443.3<br>(209.3-3,011.1) |
| **Vaccine Efficacy 70%** |  |  |  |  |  |  |  |  |  |
| 50% coverage,<br>vaccinating on day 1 | 165.8<br>(162.2-182.1) | 140.9<br>(135.4-146.7) | 29.2<br>(28.1-30.4) | 5.2<br>(3.9-5.9) | 2,526.4<br>(2,426.7-2,630.4) | 162.2<br>(152.8-172.8) | 41.4<br>(31.4-47.5) | 535.9<br>(482.6-594.9) | 1,700.3<br>(987.6-2,752.3) |
| 75% coverage,<br>vaccinating on day 1 | 79.6<br>(75.0-85.8) | 69.8<br>(64.9-74.4) | 14.5<br>(13.5-15.4) | 2.5<br>(2.0-2.9) | 1,252.1<br>(1,163.7-1,333.1) | 80.3<br>(73.8-87.0) | 20.3<br>(15.7-23.6) | 260.3<br>(233.3-291.0) | 831.0<br>(483.2-1,354.5) |
| **Vaccine Efficacy 80%** |  |  |  |  |  |  |  |  |  |
| 50% coverage,<br>vaccinating on day 1 | 147.7<br>(143.8-165.9) | 123.3<br>(118.2-128.4) | 25.6<br>(24.5-26.6) | 4.5<br>(3.4-5.1) | 2,210.3<br>(2,119.0-2,302.8) | 142.1<br>(133.1-150.8) | 36.1<br>(27.7-41.3) | 469.2<br>(424.1-522.1) | 1,451.1<br>(848.6-2,420.7) |
| 75% coverage,<br>vaccinating on day 1 | 39.3<br>(5.6-46.2) | 33.3<br>(4.7-40.5) | 6.9<br>(1.0-8.4) | 1.1<br>(0.2-1.6) | 596.7<br>(84.5-726.9) | 38.2<br>(5.4-46.9) | 9.1<br>(1.4-12.7) | 119.7<br>(17.2-154.3) | 329.1<br>(54.3-652.7) |

| Vaccine Efficacy 100% |  |  |  |  |  |  |  |  |  |
| --- | --- | --- | --- | --- | --- | --- | --- | --- | --- |
| 50% coverage,<br>vaccinating on day 1 | 107.8<br>(103.9-115.9) | 86.2<br>(82.3-90.2) | 17.9<br>(17.1-18.7) | 3.2<br>(2.4-3.6) | 1,545.7<br>(1,475.0-1,617.3) | 99.3<br>(93.0-105.8) | 25.2<br>(19.4-28.9) | 325.2<br>(293.5-363.9) | 1,031.9<br>(611.4-1,700.5) |
| 75% coverage,<br>vaccinating on day 90 | 6.5<br>(0.5-223.6) | 5.4<br>(0.4-186.2) | 1.1<br>(0.1-38.6) | 0.2<br>(0.016-6.4) | 97.0<br>(7.7-3,338.4) | 6.2<br>(0.5-208.6) | 1.5<br>(0.1-51.3) | 20.1<br>(1.6-705.5) | 60.4<br>(4.4-2,237.9) |

### 20% Vaccine Efficacy

Even when R_0_ was 2.5, which is on the lower range reported, a 20% vaccine efficacy to prevent infection was not enough to substantially reduce the epidemic, even with 100% coverage. While reducing the attack rate (to 46%), there were still a total of 143.4 million (95% UI: 134.0–152.5 million) symptomatic cases and 29.7 million (95% UI: 27.8–31.6 million) hospitalizations.

### 40% Vaccine Efficacy

When the R_0_ was 2.5, a 40% efficacious vaccine that prevented infection reduced the number of cases at the peak to 2.3 million with 100% coverage (those vaccinated and those otherwise immune) and vaccinating within 90 days of the epidemic start (Figure 1a). When covering 100% of the population, vaccination resulted in 51.6 million (95% UI: 25.3–64.5 million) symptomatic cases. A 50% coverage reduced the number of cases at the peak by 59% but still resulted in a total of 154.5 million (95% UI: 147.5–161.9 million) symptomatic cases. Vaccinating on day 180 had no impact. When R_0_ was 3.5 (Figure 2a), a 40% efficacious vaccine reduced the number of cases at the peak by 67–83% with a coverage of 75% to 100% (Table 1).

If only reducing viral shedding (Appendix Figure 1a), a 40% vaccine efficacy resulted in a total of 122.5 million (95% UI: 118.2–126.9 million) symptomatic cases even with 100% coverage.

### 60% Vaccine Efficacy

A 60% efficacious vaccine that prevented infection decreased the number of cases at the peak by 85–99% and stretch out the epidemic duration by ≥2 months when R_0_ was 2.5–3.5 with coverages ≥75% (Figures 1b and Figure 2b). For example, when R_0_ was 2.5, vaccination with 100% coverage on day 1 resulted in 2,449 (95% UI: 2,151–2,819) symptomatic cases, 89 (95% UI: 67–109) patients requiring a ventilator, and 2,815 (95% UI: 2,459–3,226) hospital bed days, costing $9.4 million in direct medical costs and $30.2 million in productivity losses. A 50% population coverage reduced the number of cases at the peak to 10.7 million (vs. 45.0 million), resulting in 115.5 million symptomatic cases (Table 1). The impact was robust to changes in the timing of vaccination, such that vaccinating on day 90 shifted the curve earlier compared to vaccinating on day 1 but still resulted in a similar magnitude (Figure 2a). However, when vaccinating on day 180, near the peak, vaccination with 100% coverage served to shorten the epidemic duration (Figure 1b). When R_0_ was 3.5, vaccination with a 75% coverage (those vaccinated and otherwise immune) resulted in 103.1 million (95% UI: 95.6–78.9 million) symptomatic cases during the epidemic. With 100% coverage, vaccination reduced the number of cases at the peak to 1.4 million cases and resulted in an 11% overall attack rate and a total of 34.4 million symptomatic cases when vaccinating on day 90 (Figure 2b; Table 1).

A vaccine that reduced viral shedding was not enough to effectively control the epidemic, even when coverage was 100% (Appendix Figure 1b). While reducing the number of cases at the peak to 20.8 million, there were still a total of 72.6 million symptomatic cases (R_0_ 2.5).

### 70% Vaccine Efficacy

When preventing infection, vaccinating with a 50% coverage on day 1 reduced the number of cases at the peak by 83%. A 60% coverage on day 1 largely extinguished the epidemic, reducing the number of cases at the peak by 93% (to 3.2 million infections), resulting in a 22% attack rate throughout the epidemic (Figure 2c) and 6.1 million (95% UI: 51.2–68.1 million) symptomatic cases, costing $938.3 billion (95% UI: $633.3-$1,432 billion) in total costs. A 60% coverage when vaccinating on day 90, resulted in a 23% attack rate over the epidemic duration and daily cases did not exceed 3.4 million. When at least 75% of the population was covered on day 1, vaccination prevented the epidemic and resulted in a total of 90,760 (95% UI: 32,544–647,422) symptomatic cases, costing $1.4 billion (95% UI: $0.5-$11.0 billion) in total costs (R_0_ 2.5). When covering at least 75% of the population on day 90, vaccination largely prevented the epidemic, resulting in an overall 5% attack rate and 13.0 million symptomatic cases (Figure 2c; Table 1). When R_0_ was 3.5, covering everyone prevented the epidemic and covering 75% of the population (those vaccinated and otherwise immune) reduced the number of cases at the peak by 93%, shifting it later into the year by 10 months when vaccinating on day 1 (Figure 2c; Table 1). With a 50% coverage, vaccinating on day 90 decreased the number of cases at the peak by 69%.

If reducing viral shedding, vaccination reduced the number of cases at the peak by 63%, resulting in 50.1 million symptomatic cases over the epidemic duration with 100% coverage (R_0_ 2.5; Appendix Figure 1c).

### 80% Vaccine Efficacy

With an 80% efficacy, a vaccine that prevents infection was reduced the number of cases at the peak by ≥36.6 million SARS-CoV-2 infections and shifted it to later in the year by more than 11 months when R_0_ was 2.5 with coverages of at least 50% and vaccinating within 90 days of the epidemic start (Figure 1d; Table 1). Covering 75% of the population resulted in a total of 3,346 (95% UI: 2,946–3,899) symptomatic cases and 3,848 (95% UI: 3,342–4,486) hospital bed days, costing $12.7 million in direct medical costs and $40.1 million in productivity losses. When R_0_ was 3.5, 50% coverage of the population reduced the number of cases at the peak by 76% but still resulted in 123.3 million symptomatic cases during the epidemic (Figure 2d, vaccinating at epidemic start). A 75% coverage decreased the total attack rate to 12%, resulting in 33.3 million symptomatic cases during the epidemic when vaccinating on day 90.

If the vaccine only reduced viral shedding, a vaccine efficacy of 80% with 100% coverage reduced the number of cases at the peak by 32.4 million (R_0_ 2.5), but still resulted in 30.6 million symptomatic cases, when vaccinating within 90 days of the epidemic start (Appendix Figure 1d; Appendix Table 2).

### 100% Vaccine Efficacy

A 100% efficacious vaccine reduced the epidemic peak by ≥44.1 million infections and shifted it ≥12 months later, even if vaccination occurred 90 after the epidemic started and coverage was 50% (R_0_ 2.5). With a 50% coverage, vaccinating resulted in 33.7 million symptomatic cases and 7.0 million hospitalizations, costing $538.1 billion in total costs during the epidemic. A 75% coverage at the start of the epidemic resulted in 928 (95% UI: 904–953) symptomatic cases and 1,067 (1,014–1,130) hospital bed days, costing $14.70 million (95% UI: $10.2-$22.0 million) in total costs. When R_0_ was 3.5, covering 50% of the population decreased the number of cases at the peak by 87%, resulting in 86.2 million symptomatic cases, even when vaccinating 90 days into the epidemic. A 75% coverage prevented the epidemic with 5.4 million symptomatic cases (2% attack rate) when vaccinating on day 90, and 246 (95% UI: 232–263) symptomatic cases when vaccinating on day 1.

If the vaccine only reduced viral shedding, covering 100% of the population still resulted in 122.9 million SARS-CoV-2 infections when R_0_ was 2.5 and 239.8 million SARS-CoV-2 infections when R_0_ was 3.5 during the epidemic.

## Discussion

Our study found that to either prevent or largely extinguish an epidemic without any other measures (e.g., social distancing), the vaccine has to have an efficacy (i.e., probability of preventing infection) of at least 70% when vaccination within 90 days of the epidemic start and covers at least 60% of the population. A 60% coverage would not be unreasonable since a recent poll by Reuters/Ipsos of 1,215 American adults found that 75% of respondents would get a coronavirus vaccine if assured it was safe.^15^ The coverage threshold, which includes those people who are vaccinated and those people who are immune for other reasons, rises the later vaccination occurs, until the threshold reaches 100% at the peak of the epidemic. This is increase is due to the fact that as the epidemic processed the prevalence of infected people is greater and greater.

There are circumstances that a vaccine with an efficacy between 40% and 70% can obviate the need for other measures but these require that 75% or more of the population be vaccinated. For example, when the vaccine efficacy is 60% at least 75% of the population would need to be vaccinated. If the R_0_ of the COVID-19 coronavirus goes from 2.5 to 3.5, then this coverage threshold would increase to 100%.

Our study focused on identifying the efficacy thresholds required to eliminate the need for other measures (e.g., social distancing) in order for life to “return to normal”, because that is a main concern of the general public.^16–20^ A vaccine alone may not be able to do this. Nonetheless, a vaccine with an efficacies lower than the thresholds we identified, could still be useful (e.g., a vaccine efficacy of 40% could prevent ≥2.8 million patients from requiring a ventilator and ≥89.5 million hospital bed days).

### Limitations

All models, by definition, are simplifications of real-life and cannot account for every possible outcome.^29^ Our model inputs drew from various sources, and new data on SARS-CoV2 continues to emerge. The course of an actual COVID-19 coronavirus epidemic may not conform to our model data and assumptions. For example, we assumed individuals mixed equally with each other, and that individuals would be vaccinated by a certain day during the epidemic; however, in reality, individuals may be vaccinated at different time points over the epidemic. Moreover, assumptions about levels of herd immunity required may depend on differing individual susceptibility to SARS-CoV-2.

### Conclusions

Our study found that to either prevent or largely extinguish an epidemic without any other measures (e.g., social distancing), the vaccine has to have an efficacy of at least 70% when vaccination occurs within 90 days of the start of the epidemic and covers at least 60% of the population. This efficacy threshold increases the deeper one goes into the pandemic. A vaccine with an efficacy between 40% and 70% can still obviate the need for other measures under certain circumstances such as much higher, and potentially unachievable, vaccination coverages.

## Data Availability

All relevant data available in manuscript.

## Appendix

**Appendix Table 1.** Model parameter inputs, values, and sources

| Parameter | Mean or Median | Standard Error<br>or Range | Source |
| --- | --- | --- | --- |
| **SARS-CoV2 Transmission** |  |  |  |
| Incubation period (days) | 5.2 | 4.1 – 7.0 | 12 |
| Infectious period (days) |  | 3 - 14 | 21-24 |
| **Costs (2020 US\$)** | | | |
| Annual wages (all occupations) | 40,993 | 21,950 - 104,403<sup>a</sup> | 9 |
| Ambulatory care visit |  | 110.43 - 148.33 | 25 |
| Over the counter medications, daily |  |  |  |
| 0-12 years old<sup>b</sup> | 3.87 | 2.10 | 26 |
| ≥13 years old<sup>c</sup> | 0.46 | 0.17 | 26 |
| Hospitalization for pneumonia<sup>d</sup> |  |  |  |
| 0-17 years old | 12,502.30 | 1,508.04 | 27 |
| 18-44 years old | 10,627.15 | 1,045.06 | 27 |
| 45-64 years old | 13,718.14 | 1,238.76 | 27 |
| 65-84 years old | 12,264.39 | 478.40 | 27 |
| ≥85 years old | 10,982.73 | 518.29 | 27 |
| Hospitalization for severe non-pneumonia (all ages)<sup>e</sup> | 6,886.53 | 1,182.99 | 27 |
| Hospitalization for sepsis<sup>f</sup> |  |  |  |
| 0-17 years old<sup>g</sup> | 22,694.30 | 1,861.33 | 27 |
| 18-44 years old | 43,778.39 | 5,382.40 | 27 |
| 45-64 years old | 38,734.24 | 2,725.10 | 27 |
| 65-84 years old | 30,308.29 | 1,367.91 | 27 |
| ≥85 years old | 22,694.30 | 1,861.33 | 27 |
| Hospitalization for ARDS<sup>h</sup> |  |  |  |
| 0-17 years old | 42,350.58 | 4,198.97 | 27 |
| 18-44 years old | 26,210.96 | 1,558.61 | 27 |
| 45-64 years old | 19,863.98 | 453.92 | 27 |
| 65-84 years old | 18,718.55 | 335.69 | 27 |
| ≥85 years old | 16,559.75 | 754.12 | 27 |
| **Probabilities** |  |  |  |
| Asymptomatic infection | 0.179 | 0.155 - 0.202 | 28 |
| Missing work/school | 1.0 |  | Assumption |
| Ambulatory care |  |  |  |
| 0-4 years old | 0.455 | 0.098 | 29 |
| 5-17 years old | 0.318 | 0.061 | 29 |
| 18-64 years old | 0.313 | 0.014 | 29 |
| ≥65 years old | 0.62 | 0.027 | 29 |
| Probability of hospitalization, given infection |  |  |  |
| 0-19 years old | 0.016 |  | 11 |
| 20-44 years old | 0.143 |  | 11 |
| 45-64 years old | 0.208 |  | 11 |
| 65-84 years old | 0.292 |  | 11 |
| ≥85 years old | 0.313 |  | 11 |
| Probability of ICU admission |  |  |  |
| 0-19 years old | 0.0 |  | 11 |
| 20-44 years old | 0.1399 |  | 11 |
| 45-64 years old | 0.2422 |  | 11 |
| 65-84 years old | 0.3048 |  | 11 |
| ≥85 years old | 0.2013 |  | 11 |
| Probability of Mortality |  |  |  |
| 0-19 years old | 0.0 |  | 11 |
| 20-44 years old | 0.007 |  | 11 |
| 45-64 years old | 0.0456 |  | 11 |
| 65-84 years old | 0.1109 |  | 11 |
| ≥85 years old | 0.3323 |  | 11 |
| Pneumonia, given hospitalization | 0.79 | 0.711-.869<sup>ii</sup> | 30 |
| ARDS, requiring ventilator use in ICU | 0.73 | 0.1697 | 31, 32 |
| Age-group, given infection |  |  |  |
| 0-19 years old | 0.0502 |  | 11 |
| 20-44 years old | 0.2879 |  | 11 |
| 45-64 years old | 0.3503 |  | 11 |
| 65-84 years old | 0.2528 |  | 11 |
| ≥85 years old | 0.0588 |  | 11 |
| **Durations (days)** |  |  |  |
| Ambulatory care | 0.5 |  | Assumption |
| Telephone consult (tele-med; minutes) | 17 | 10 - 24<sup>j</sup> | 33 |
| Duration of symptoms with mild illness | 7 | 3-17 | 23, 34, 35 |
| Duration of symptoms prior to hospital admission | 7 | 3-9<sup>j</sup> | 31, 36 |
| Hospitalization for pneumonia<sup>d</sup> |  |  |  |
| 0-17 years old | 4.7 | 0.4 | 27 |
| 18-44 years old | 4.3 | 0.4 | 27 |
| 45-64 years old | 5.1 | 0.2 | 27 |
| 65-84 years old | 5.5 | 0.2 | 27 |
| ≥85 years old | 5.0 | 0.2 | 27 |
| Hospitalization for severe non-pneumonia (all ages)<sup>e</sup> | 3.1 | 0.5 | 27 |
| Hospitalization for sepsis<sup>f</sup> |  |  |  |
| 0-17 years old<sup>g</sup> | 7.3 | 0.5 | 27 |
| 18-44 years old | 11.2 | 1.3 | 27 |
| 45-64 years old | 10.7 | 0.5 | 27 |
| 65-84 years old | 8.8 | 0.4 | 27 |
| ≥85 years old | 7.3 | 0.5 | 27 |
| Hospitalization for ARDS<sup>h</sup> |  |  |  |
| 0-17 years old | 9.5 | 0.75 | 27 |
| 18-44 years old | 8.8 | 0.5 | 27 |
| 45-64 years old | 7.1 | 0.1 | 27 |
| 65-84 years old | 7.0 | 0.1 | 27 |
| ≥85 years old | 6.1 | 0.3 | 27 |
| **Utility weights** |  |  |  |
| Healthy QALY |  |  |  |
| <17 years old | 1 | - | 37 |
| 18-64 years old | 0.92 | - | 37 |
| ≥65 years old | 0.84 | - | 37 |
| Mild non-specific symptoms<sup>k</sup> | 0.648 | 0.103 | 38-47 |
| Hospitalized, non-pneumonia symptoms<sup>i</sup> | 0.514 | 0.089 | 40, 47, 48 |
| Pneumonia | 0.496 | 0.17 | 49-53 |
| Sepsis | 0.467 | 0.18 | 53-59 |
| ARDS | 0.10 | 0.08-0.15 | 60 |

<sup>a</sup>Values are 95% confidence interval
<sup>b</sup>Assumes 5 to 10 mg/kg orally every 6 to 8 hours as needed OR 10 to 15 mg/kg orally every 4 to 6 hours as needed
<sup>c</sup>Assumes 200 mg orally every 4 to 6 hours as needed
<sup>d</sup>Uses International Classification of Diseases, Tenth Revision, Clinical Modification (ICD10) code #J13 Pneumonia due to *Streptococcus pneumoniae*
<sup>e</sup>Uses International Classification of Diseases, Tenth Revision, Clinical Modification (ICD10) code #J11.89 Influenza due to unidentified influenza virus with other manifestations
<sup>f</sup>Uses International Classification of Diseases, Tenth Revision, Clinical Modification (ICD10) code #R65.21 Severe sepsis with septic shock
<sup>g</sup>Data for age-group unavailable and uses lowest values of all age-groups as a proxy
<sup>h</sup>Uses International Classification of Diseases, Tenth Revision, Clinical Modification (ICD10) code #J96.22 Acute and chronic respiratory failure with hypercapnia for 18 years and older and ICD10 code #J96.20 Acute and chronic respiratory failure, unspecified whether with hypoxia or hypercapnia for 0 to 17-year olds
<sup>i</sup>Values are interquartile range
<sup>j</sup>Values are 10%-90%
<sup>k</sup>Uses influenza without hospitalization as a proxy
<sup>l</sup>Uses influenza with hospitalization as a proxy

**Appendix Figure 1.**
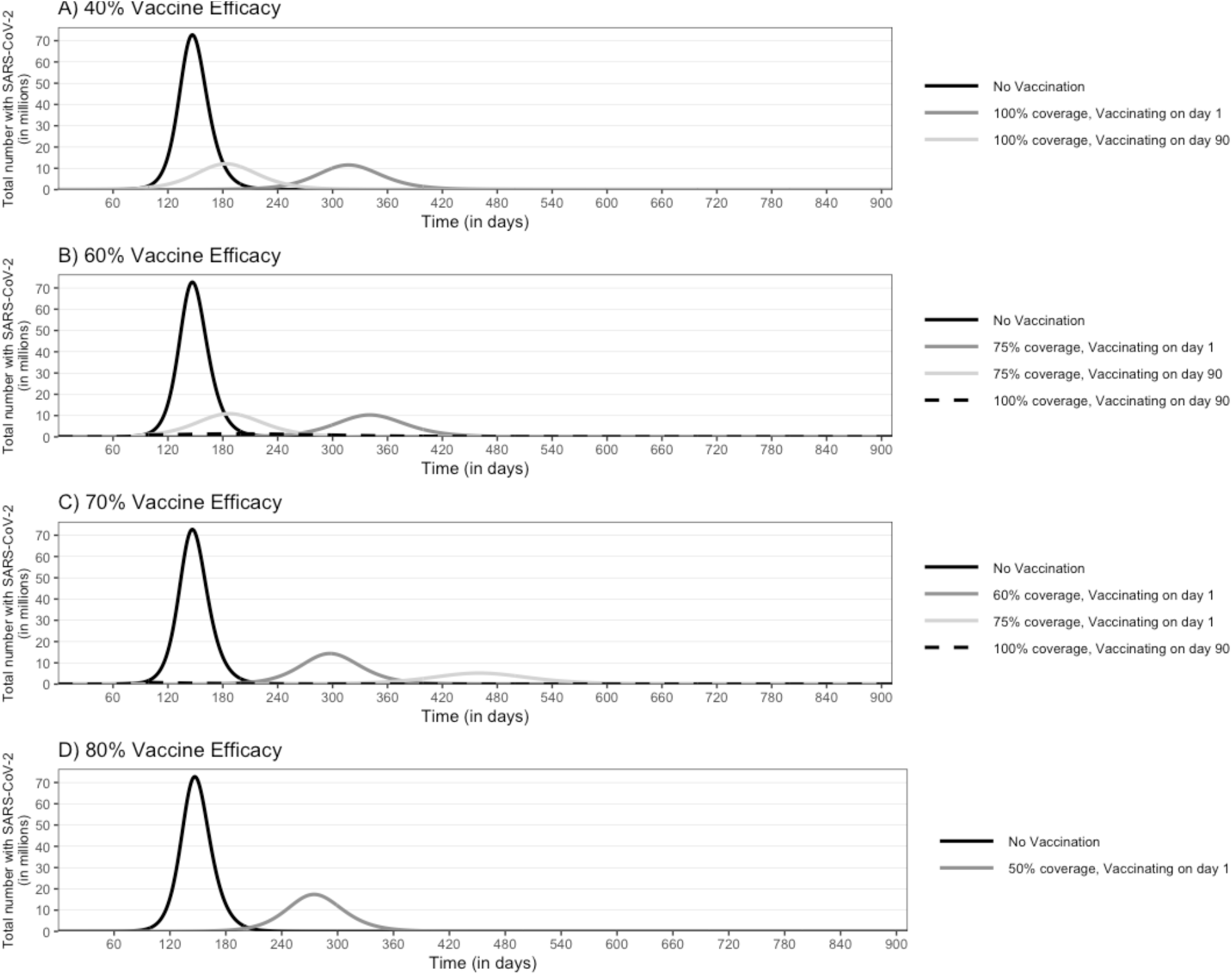
Epidemic curves for an epidemic with an R0 of 2.5 and the impact of a vaccine that reduces viral shedding implemented at different points in the epidemic with a vaccine efficacy of A) 40%, B) 60%, C) 70%, and D) 80%

**Appendix Table 2.** Number of clinical outcomes, resource use, and costs [median (95% uncertainty interval)] due to COVID-19 during the course of an epidemic and the impact of vaccination with a vaccine that reduces viral shedding, varying with vaccine efficacy

|  | Total Infected<br>with SARS-CoV-2 (in<br>millions) | Symptomatic<br>Cases<br>(in millions) | Hospitalized<br>(in millions) | Number of<br>Patients<br>Ventilated<br>(in millions) | Deaths<br>(in thousands) | Total Beds<br>Days (millions) | Ventilated<br>Days<br>(millions) | Direct Medical<br>Costs (in<br>billions) | Productivity Losses<br>(in billions) |
| --- | --- | --- | --- | --- | --- | --- | --- | --- | --- |
| **R0 of 2.5** |  |  |  |  |  |  |  |  |  |
| No Vaccination | 282.5<br>(280.7-284.3) | 232.1<br>(223.9-240.0) | 48.1<br>(46.4-49.8) | 8.5<br>(6.5-9.7) | 4,160.9<br>(4,014.2-4,302.0) | 267.2<br>(251.7-282.6) | 68.0<br>(51.8-77.9) | 883.5<br>(808.5-980.8) | 2,796.4<br>(1,661.8-4,515.5) |
| **Vaccine Efficacy 60%** |  |  |  |  |  |  |  |  |  |
| 100% coverage,<br>vaccinating on day 1 | 228.1<br>(220.2-244.8) | 72.7<br>(70.0-75.3) | 15.1<br>(14.5-15.6) | 2.7<br>(2.0-3.0) | 1,302.6<br>(1,255.1-1,350.1) | 83.7<br>(78.6-88.6) | 21.2<br>(16.3-24.4) | 276.0<br>(250.8-305.0) | 858.9<br>(504.3-1,410.2) |
| **Vaccine Efficacy 70%** |  |  |  |  |  |  |  |  |  |
| 75% coverage,<br>vaccinating on day 1 | 494.4<br>(447.2-497.8) | 90.8<br>(87.6-94.2) | 18.8<br>(18.2-19.5) | 3.3<br>(2.5-3.8) | 1,627.8<br>(1,569.9-1,688.6) | 104.7<br>(98.6-111.2) | 26.7<br>(20.4-30.5) | 345.8<br>(313.4-382.8) | 1,070.4<br>(633.2-1,787.4) |
| 100% coverage,<br>vaccinating on day 1 | 480.7<br>(372.9-511.3) | 50.1<br>(48.4-52.0) | 10.4<br>(10.0-10.8) | 1.8<br>(1.4-2.1) | 897.3<br>(867.0-932.0) | 57.7<br>(54.4-61.2) | 14.6<br>(11.3-16.7) | 189.0<br>(171.3-210.2) | 598.3<br>(346.8-962.0) |
| **Vaccine Efficacy 80%** |  |  |  |  |  |  |  |  |  |
| 75% coverage,<br>vaccinating on day 1 | 227.3<br>(220.1-245.1) | 72.7<br>(70.1-75.4) | 15.1<br>(14.5-15.6) | 2.7<br>(2.1-3.0) | 1,302.6<br>(1,256.5-1,350.9) | 83.8<br>(79.1-88.9) | 21.4<br>(16.4-24.4) | 276.3<br>(251.3-304.9) | 874.1<br>(520.8-1,404.4) |
| 100% coverage,<br>vaccinating on day 1 | 186.1<br>(181.4-194.5) | 29.8<br>(28.7-30.9) | 6.2<br>(5.9-6.4) | 1.1<br>(0.8-1.2) | 533.7<br>(514.4-553.9) | 34.3<br>(32.3-36.3) | 8.8<br>(6.8-10.0) | 111.6<br>(101.6-123.6) | 348.9<br>(209.5-566.6) |
| 100% coverage,<br>vaccinating on day 90 | 188.9<br>(182.6-204.6) | 30.5<br>(28.9-50.8) | 6.3<br>(6.0-10.5) | 1.1<br>(0.9-1.8) | 546.4<br>(518.8-909.9) | 35.2<br>(32.5-57.6) | 9.0<br>(7.0-14.7) | 116.8<br>(104.4-194.6) | 379.0<br>(219.8-658.8) |
| **Vaccine Efficacy 100%** |  |  |  |  |  |  |  |  |  |
| 75% coverage,<br>vaccinating on day 1 | 198.8<br>(192.7-208.4) | 39.6<br>(38.2-41.1) | 8.2<br>(7.9-8.5) | 1.4<br>(1.1-1.7) | 710.7<br>(684.6-737.0) | 45.6<br>(42.9-48.4) | 11.6<br>(8.9-13.3) | 149.3<br>(135.3-165.8) | 453.6<br>(269.2-746.9) |
| 100% coverage,<br>vaccinating on day 90 | 127.7<br>(124.5-168.4) | 0.6<br>(0.1-20.0) | 0.12<br>(0.018-4.1) | 0.02<br>(0.003-0.70) | 9.9<br>(1.6-358.8) | 0.6<br>(0.1-22.9) | 0.2<br>(0.025-5.6) | 2.1<br>(0.3-80.5) | 6.5<br>(0.9-234.5) |
| **R0 of 3.5** |  |  |  |  |  |  |  |  |  |
| No Vaccination | 312.9 | 257.1 | 53.3 | 9.5 | 4,608.3 | 296.0 | 76.2 | 978.9 | 3,141.3 |
|  | (311.9-314.0) | (248.7-264.8) | (51.6-54.9) | (7.2-10.7) | (4,458.0-4,747.6) | (281.4-312.1) | (57.6-85.7) | (896.4-1,082.3) | (1,871.3-5,059.2) |
| **Vaccine Efficacy 60%** |  |  |  |  |  |  |  |  |  |
| 100% coverage,<br>vaccinating on day 1 | 285.4<br>(284.3-288.1) | 93.8<br>(90.7-96.6) | 19.4<br>(18.8-20.0) | 3.4<br>(2.6-3.9) | 1,681.5<br>(1,626.6-1,732.2) | 107.9<br>(102.5-113.8) | 27.4<br>(21.0-31.3) | 357.4<br>(324.1-396.0) | 1,122.7<br>(648.1-1,835.6) |
| **Vaccine Efficacy 70%** |  |  |  |  |  |  |  |  |  |
| 100% coverage,<br>vaccinating on day 1 | 692.6<br>(688.6-695.7) | 68.1<br>(65.9-70.2) | 14.1<br>(13.7-14.5) | 2.5<br>(1.9-2.8) | 1,220.2<br>(1,180.6-1,258.0) | 78.3<br>(74.2-82.6) | 19.9<br>(15.0-22.7) | 258.9<br>(235.4-285.2) | 803.4<br>(471.8-1,328.1) |
| **Vaccine Efficacy 80%** |  |  |  |  |  |  |  |  |  |
| 100% coverage,<br>vaccinating on day 1 | 266.1<br>(264.9-289.0) | 43.5<br>(42.2-45.0) | 9.0<br>(8.7-9.3) | 1.6<br>(1.2-1.8) | 780.6<br>(755.9-806.3) | 50.1<br>(47.3-52.8) | 12.7<br>(9.7-14.6) | 166.0<br>(151.7-183.2) | 517.1<br>(305.5-831.1) |
| **Vaccine Efficacy 100%** |  |  |  |  |  |  |  |  |  |
| 75% coverage,<br>vaccinating on day 1 | 271.7<br>(270.6-286.5) | 55.6<br>(54.0-57.4) | 11.5<br>(11.2-11.9) | 2.0<br>(1.6-2.3) | 996.8<br>(967.5-1,028.5) | 64.1<br>(60.4-67.4) | 16.4<br>(12.5-18.6) | 212.4<br>(192.0-232.2) | 667.5<br>(396.3-1,094.4) |

